# Early developmental milestone clusters of autistic children based on electronic health records

**DOI:** 10.1101/2024.01.16.24301373

**Authors:** Ayelet Ben-Sasson, Joshua Guedalia, Keren Ilan, Galit Shefer, Roe Cohen, Lidia V Gabis

## Abstract

ASD children vary in symptoms, co-morbidities and response to interventions. This study aimed to identify clusters of ASD children with a distinct pattern of attaining early developmental milestones. Clustering of 5,836 ASD children was based on attainment of 43 gross motor, fine motor, language and social developmental milestones in the first three years of life as recorded in baby wellness visits.

K-means cluster analysis detected 4 Early Developmental Milestone (EDM) clusters: typical (n=1,686); mild (n=1,691); moderate (n=2,265); and severe (n=194). Most prominent cluster differences were in the language domain. The severe cluster showed earlier and greater developmental delay across domains, unique early gross motor delays, and more were born preterm via cesarian section. Moderate cluster had poor language development prominently in the second year of life, later fine motor delays. Mild cluster had language delays in the third year of life. The typical cluster mostly passed milestones. EDM clusters differed demographically, with higher socioeconomic status in typical cluster and lowest in severe cluster. Moderate cluster had more immigrant and non-Jewish mothers followed by the mild cluster. The rates of parental concerns and provider developmental referrals were significantly higher in the severe, followed by the moderate, mild, and typical EDM clusters.

ASD children’s language and motor delay in the first three years can be grouped by common magnitude and onset profiles as distinct groups that may link to specific etiologies (like prematurity or genetics) and to specific intervention programs. Early ASD screening should be tailored to these different developmental profiles.

## Introduction

Autism spectrum disorder (ASD) is characterized by a range of neurodiverse impairments, marked by common social communication deficits and restricted, repetitive behaviors [1]. ASD exhibits multidimensional diversity with discernible endophenotypes, representing distinct traits that may indicate different etiologies [2,3]. These endophenotypes can inform personalized therapeutic strategies based on severity [4,5], etiology [6], gene expression [7], developmental/IQ/adaptive levels [5,8,9], comorbid conditions [10–12], and response to intervention [3,6]). ASD heterogeneity is also evident in the variability of early developmental milestones, influenced by genetics, intellect, sex, and comorbidities [7,13]. Identifying distinct developmental trajectories in ASD is crucial for designing a detection protocol that accounts for various early profiles, particularly those that may be overlooked or masked by other symptoms, as seen in girls and in children with comorbidities [14,15]. Recognizing these different trajectories is essential for tailoring early interventions to meet the unique developmental needs of each subgroup. Utilizing big data, rich in both quantity and quality, presents an opportunity for unsupervised data-driven discoveries of this variability [16].

The twofold objective of this study was to identify subgroups of ASD children who share early developmental trajectories, as documented in their routine baby wellness electronic health records (EHR) and to capture the array of domains affected (e.g., gross and fine motor skills, language, social) and their patterns of change during the first three years of life.

### Different early developmental trajectories in ASD

One barrier to early ASD detection is the variability in early behavioral markers, their onset, and progression [17,18]. ASD Children follow diverse developmental trajectories, with some experiencing persistent delays, others stabilizing and some exhibiting regression [17–19]. This variability extends to different developmental domains, further complicating the interpretation and prediction of the developmental pattern [5,20]. A wide-scale, ASD health records study including among other parameters eight milestones attained up to the age of five years, found a median delay of 0.7 to 19.7 months, with larger gaps for children with intellectual disability and genetic etiology [13]. The age of ASD diagnosis varies among children, partly due to their early developmental profile. In another health records study, 36.7% of ASD children were initially referred due to motor development delays at earlier ages. They had later and lower rates of referral for speech and language delays (28.6%), and communication delays (13%) [21]. Interestingly, not achieving more milestones may delay ASD diagnosis [22], as multiple missed milestones may result in a diagnosis of global developmental delay, which might imply a differential diagnosis, and as per the DSM-5, diagnosing ASD alongside developmental delay requires demonstrating the severity of ASD symptoms [23].

Two studies, employing a developmental checkup protocol similar to ours, explored the early developmental profiles of ASD children [22,24]. At the age of 9 months, ASD children exhibited distinct developmental trajectories, particularly in the motor and social communication domains, when compared to children with other disorders [24]. The number of unmet developmental milestones was found to be associated with increased ASD likelihood, and this association grew stronger with age. Failure in attaining motor and social milestones were consistent indicators of ASD across ages, while language delays became noticeable after 12 months. The most robust prediction of ASD was observed for social milestone failures at the 9–12 month visit and motor milestone failures at the 12–18 month visit, irrespective of sex and ethnicity [22]. The present study expands upon this research by examining developmental variability within the ASD phenotype during the first three years of life.

Most efforts to unravel the heterogeneity of ASD have primarily focused on cohorts diagnosed later, mainly relying on developmental level scores [9,25,26], rather than examining early developmental trajectories. Preschool studies have revealed that ASD subgroups vary in the severity of their ASD symptoms, particularly in the social communication domain, IQ, and adaptive scores, but not significantly in their motor scores [5]. Other research with preschool-aged children has identified three distinct combinations: those with high adaptive levels and low ASD severity, those with high adaptive levels and high ASD severity, and those with low adaptive levels and high ASD severity [9]. In a study involving 707 ASD children ages 2–5 years, 4 subgroups emerged: 1. General developmental delay with moderate-to-high ASD severity (34%), 2. Mild language delay coupled with cognitive rigidity (28%), 3. Significant developmental delay with repetitive motor behaviors (26%), and 4. Mild language and motor delays with dysregulation (12%). These findings underscore the importance of considering factors beyond early language development, as at least two clusters exhibited mild language delays with cognitive rigidity. Moreover, clusters with early motor and language delays highlight the need for closer monitoring and additional screening during the early years of life [5].

Very few ASD clustering studies include developmental milestones in their classification [5,13], mainly due to lack of prospective data preceding the autism diagnosis. Classification research aiming to investigate group differences in developmental change over time across multiple dimensions requires a very large ASD dataset, which is possible through health record research.

Studying clinical phenotypes and clustering symptoms into endophenotypes can help identify polygenic mechanisms in various complex conditions, including ASD [27].

The current study is part of a larger project analyzing the baby wellness database of the Israeli Ministry of Health (MoH) and a diagnosis of ASD in the Israeli National Social Security system. This database includes routine developmental checkups administered to the general population over 11 visits. Previous research of the distribution of milestone attainment up to 6 years of age in the MoH database of typically developing children showed that about one-third of the milestones are achieved by more than 95% of children at the earliest visits tested, and another tenth by 90% to 95% of children [28]. This study aimed to identify subgroups of ASD children, distinguished by their developmental progress during the first three years of life. Furthermore, we aimed to validate clusters by developmental referral rates and parental concerns, and to explore their unique familial (e.g., maternal age, immigration) and birth parameters (e.g., prematurity, sex, and birth weight).

## Methods

### Procedures

This study was approved by the MoH Ethics Committee (#15/2021), which waived the need for informed consent. Data were extracted on 2 May 2023 and anonymized by the TIMNA project personnel (Israeli MoH Big Data Platform). The EHR represented children born 1 January 2014 through 31 December 2019. Researchers did not have access to identifying information of participants. TIMNA personnel matched MoH EHR data with corresponding Israeli National Social Security system ASD diagnosis claim data.

The MoH is the main provider of developmental checkups at 503 clinics across Israel, covering 70% of the population. The remaining 30% are followed by 2 Health Maintenance Organizations according to the same national guidelines. The services include immunizations, growth and developmental surveillance, as well as medical and environmental risk detection for the mother and child. The services are provided by nurses with specific training in maternal and child health and a developmental pediatrician. There are 11 visits from birth to age 6 years, with guidelines based on the child’s age and risk status. Attendance is highest during the 3 first years.

### Participants

The database initially extracted consisted of 780,610 records of children born from January 2014 through December 2019, with at least 2 years of MoH baby wellness surveillance. The current study focused on records of children who had an ASD diagnosis based on their Israel National Social Security disability claim records. Diagnosis requires assessment by a psychiatrist, neurologist, or developmental pediatrician, standard intelligence/developmental test, and an ASD diagnostic tool approved by the MoH. A total of 15,956 had an ASD diagnosis. For meaningful clustering, the current study required having full milestone data up to age three years, resulting in a final sample of 5,836 (36.6%) ASD children (see supplemental figure 1; compared to 35.24% full milestone data up to age three years in typically developing in this database).

### Measures

The well-baby developmental protocol includes checkup of progress in 60 age-related milestones across gross motor, fine motor, language, and personal-social developmental domains. The milestones included are based in part on the Denver Developmental Screening Test [29]. At each of nine age groups from six weeks to six years, the nurses check 4 to 9 milestones. Performance for each milestone is recorded. During developmental checkups, parents are also prompted to report their concerns regarding development and hearing. For this study, we focused on milestones tested up to 36 months, as beyond that data were sparse.

### Data analysis

Milestone performance was re-coded as passed or failed for entering the cluster analysis. Passed was defined as performing the milestone at the checkup or a positive parent report during any visit. Failed was defined as a report of not performing the milestone. Examination of 45 milestones tested by 36 months indicated that 2 milestones had above 40% missing data in the entire 780,610 cohort and were therefore dropped from analysis.

K-means clustering, an unsupervised machine learning technique, was employed to partition the 5,836 children into 9 distinct clusters based on 43 binary milestones. The dataset consisted of 250,948 data points. The algorithm involved an iterative process of assigning data points to the closest cluster center, known as centroids, based on Euclidean distance. We selected k=9 (clusters) and then used domain knowledge to group together clusters into 4 clinically similar clusters.

The mean number of failed milestones were described for each domain and cluster. In addition, milestone percentiles were computed based on the child’s age at first performance of the milestone relative to typically developing children born at term (n=703,551). Milestone percentile was not computed for a failed performance. Mean domain percentiles per time point were computed for cases in which at least one milestone was attained within that domain. Note that mean domain percentiles per time point was missing for only 3 children (0.05%).

Clusters were compared on demographic, birth and post-natal variables, as well as developmental referrals and concerns following normality testing. Nominal variables were compared using Chi-square tests and normally distributed continuous variables using ANOVA. Pregnancy week did not distribute normally (p<.001) and therefore was compared using Kruskal-Wallis test. Main results were followed up with post hoc tests and include corresponding effect size values. Missing values were not included in these cluster comparisons.

## Results

K-means cluster analysis using k=9, yielded 9 clusters. The largest cluster included 1,686 and the smallest 194 participants (see supplemental materials figure 2 for histogram showing the distribution of cohort between clusters). Those with similar patterns of development were grouped into four Early Developmental Milestone (EDM) clusters of: typical (n=1,686), mild (n=1,691), moderate (n=2,265), and severe (n=194). Figure 1 presents the percentage of children in each cluster who passed a milestone (see supplemental figure 3 for this data by 9 and 4 clusters).

**Figure 1.**
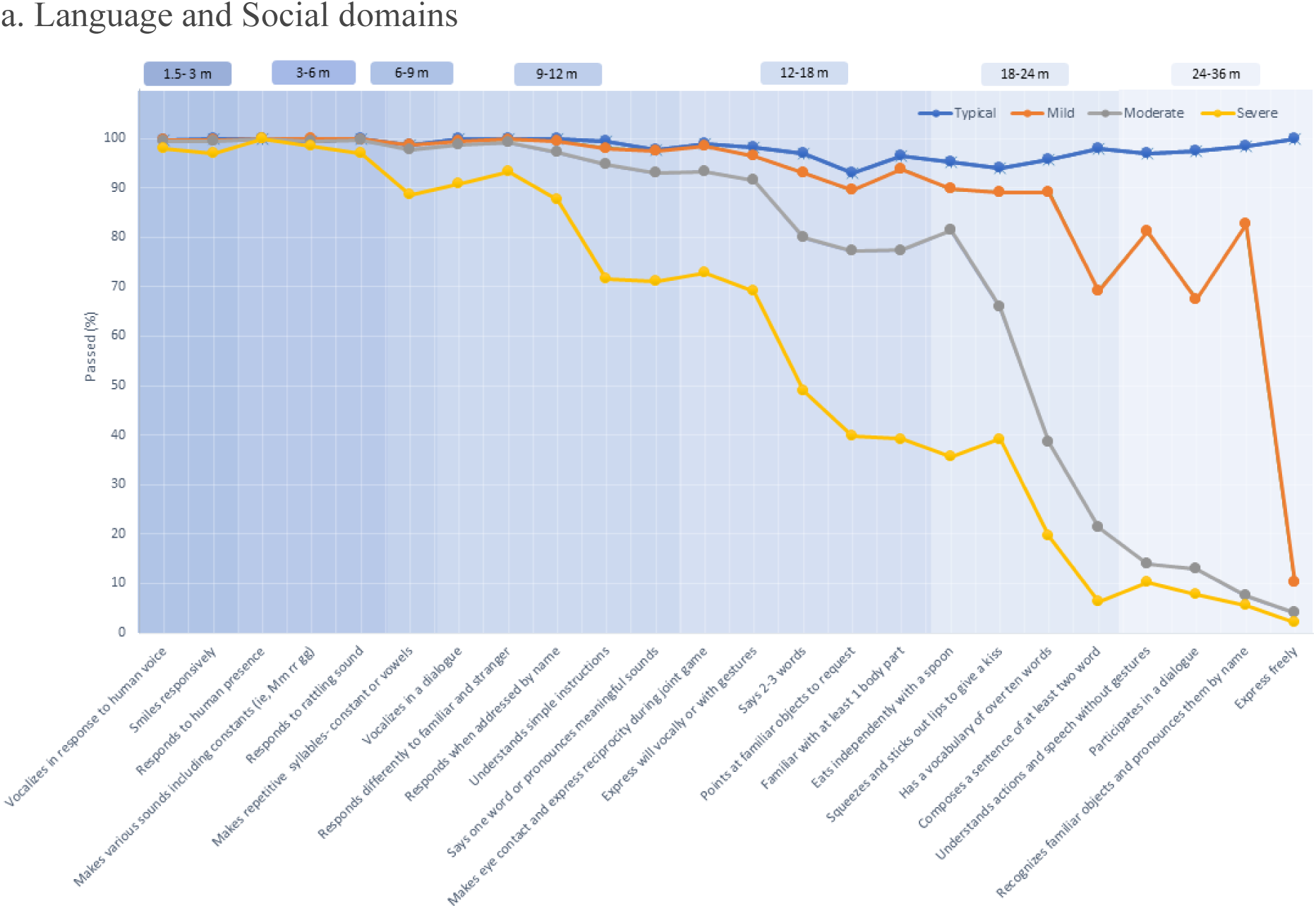
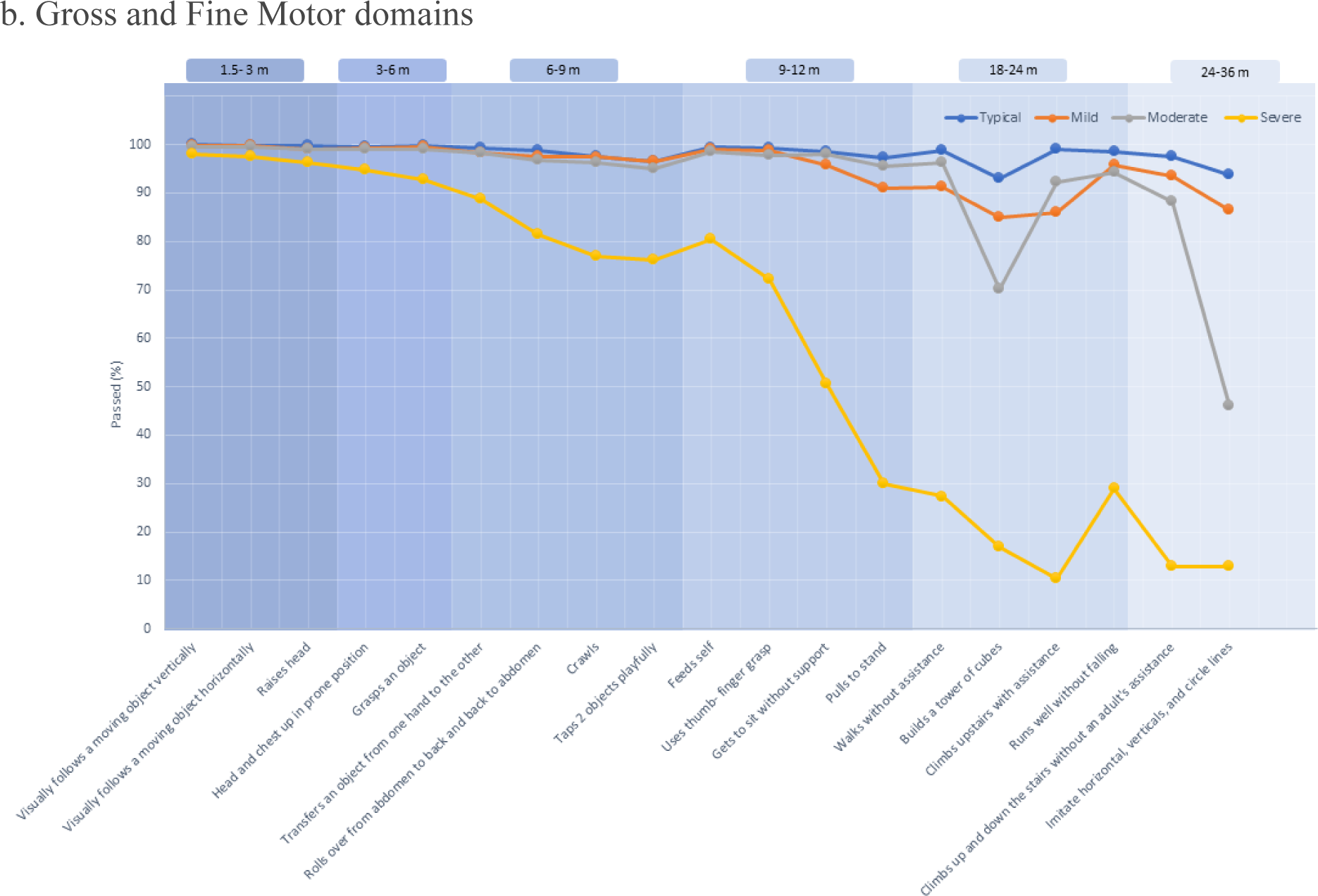
a. Percentage of those performing language and social milestones by EDM cluster. b. Percentage of those performing gross and fine motor milestones by EDM cluster.

### Demographic comparison of EDM clusters

Table 1 displays differences between EDM clusters in child and family demographic and birth parameter variables. Noteworthy are those differences with moderate-to-high effect sizes (V>.10). The mild cluster had significantly fewer Jewish families than the other clusters. The moderate cluster had significantly fewer Jewish families than the typical cluster. The severe cluster did not differ in rates from moderate or typical clusters. The mild cluster also had the highest rate of immigrant mothers, followed by the moderate group, with no differences between the severe and typical clusters. The percentage of families with high SES status was highest in the typical cluster, followed by the mild, moderate and severe EDM clusters.

**Table 1.**
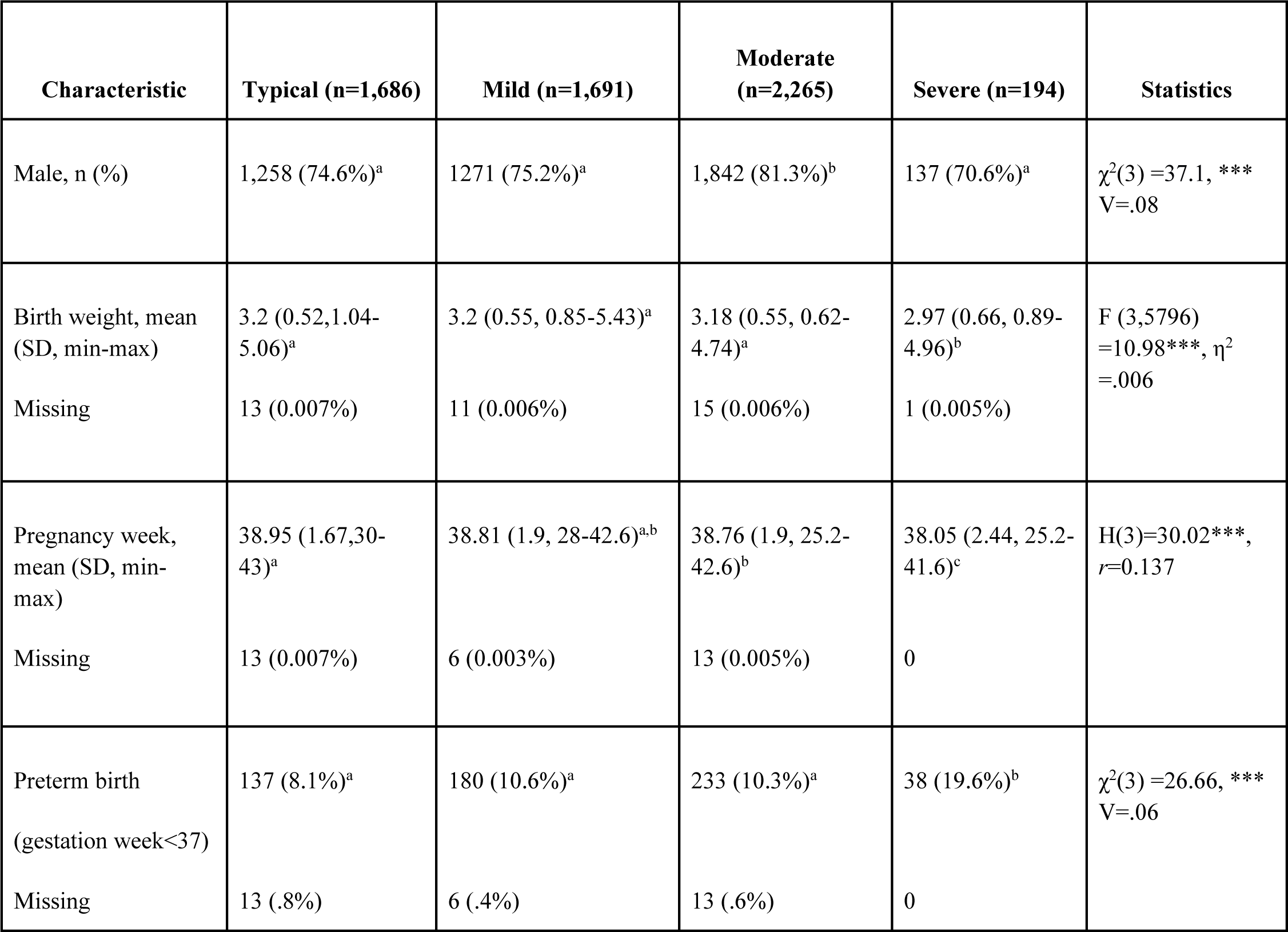
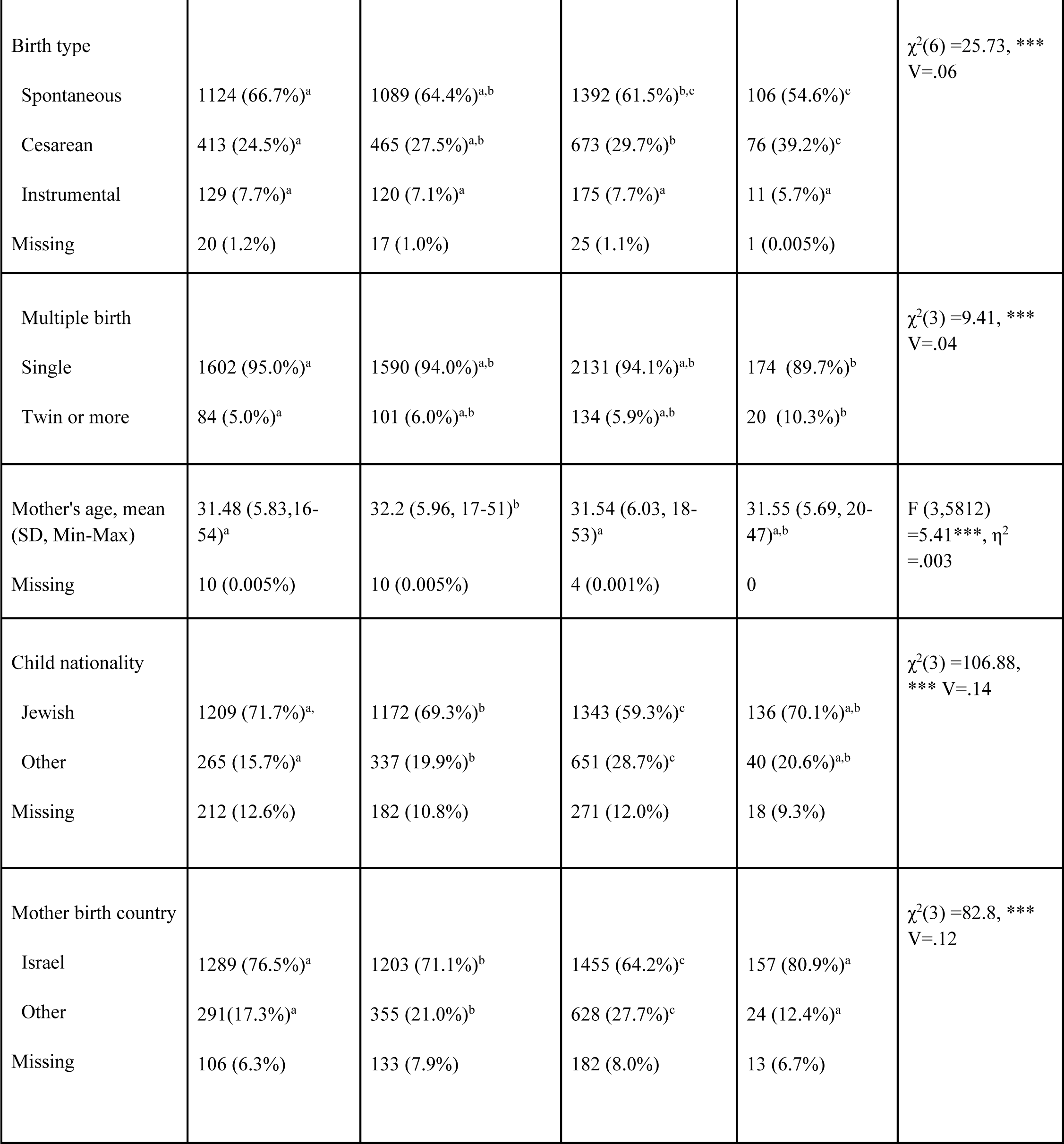
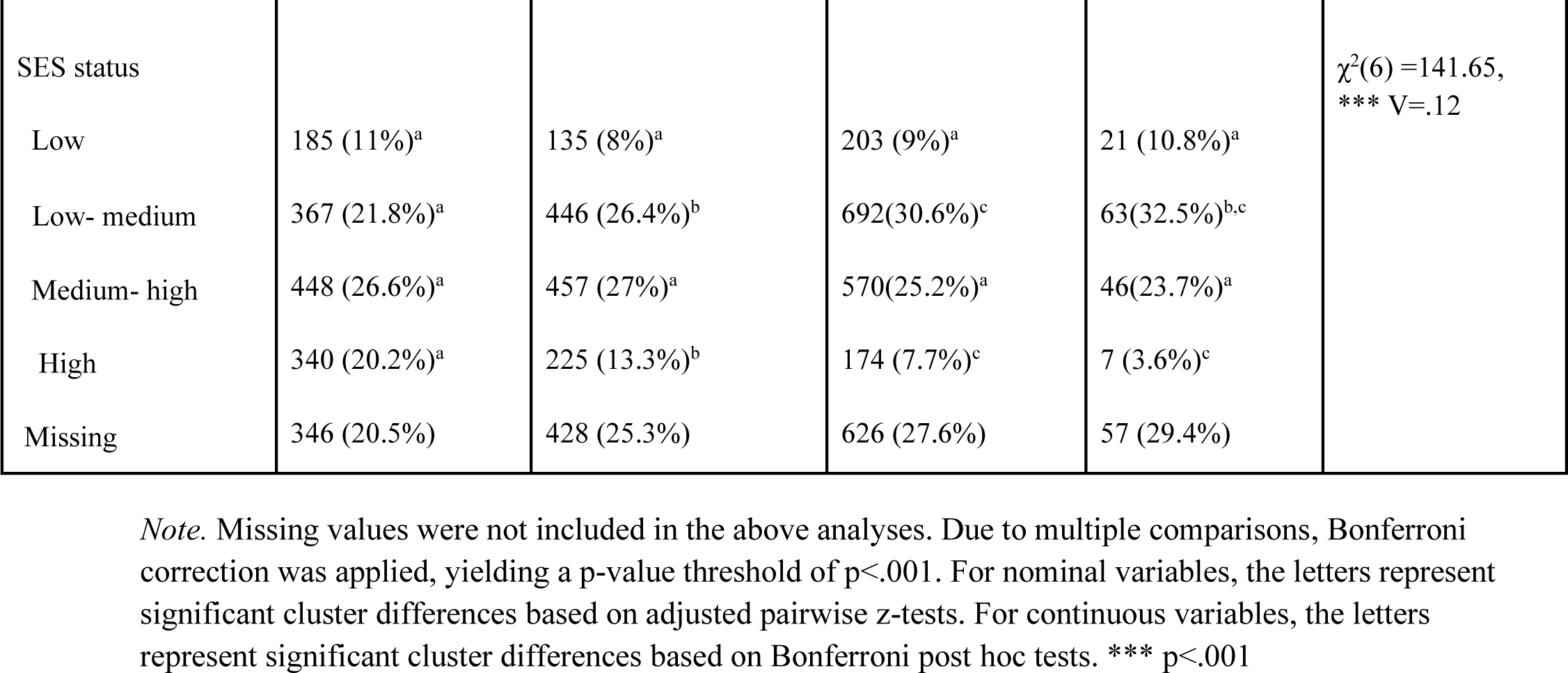
Comparison of Birth and Demographic Characteristics by EDM Cluster.

| Characteristic | Typical (n=1,686) | Mild (n=1,691) | Moderate (n=2,265) | Severe (n=194) | Statistics |
| --- | --- | --- | --- | --- | --- |
| Male, n (%) | 1,258 (74.6%)<sup>a</sup> | 1271 (75.2%)<sup>a</sup> | 1,842 (81.3%)<sup>b</sup> | 137 (70.6%)<sup>a</sup> | $\chi^2(3) = 37.1$, ***<br>V=.08 |
| Birth weight, mean (SD, min-max) | 3.2 (0.52, 1.04-5.06)<sup>a</sup> | 3.2 (0.55, 0.85-5.43)<sup>a</sup> | 3.18 (0.55, 0.62-4.74)<sup>a</sup> | 2.97 (0.66, 0.89-4.96)<sup>b</sup> | F (3,5796) = 10.98***, $\eta^2$ = .006 |
| Missing | 13 (0.007%) | 11 (0.006%) | 15 (0.006%) | 1 (0.005%) |  |
| Pregnancy week, mean (SD, min-max) | 38.95 (1.67, 30-43)<sup>a</sup> | 38.81 (1.9, 28-42.6)<sup>a,b</sup> | 38.76 (1.9, 25.2-42.6)<sup>b</sup> | 38.05 (2.44, 25.2-41.6)<sup>c</sup> | H(3)=30.02***, $r=0.137$ |
| Missing | 13 (0.007%) | 6 (0.003%) | 13 (0.005%) | 0 |  |
| Preterm birth (gestation week<37) | 137 (8.1%)<sup>a</sup> | 180 (10.6%)<sup>a</sup> | 233 (10.3%)<sup>a</sup> | 38 (19.6%)<sup>b</sup> | $\chi^2(3) = 26.66$, ***<br>V=.06 |
| Missing | 13 (.8%) | 6 (.4%) | 13 (.6%) | 0 |  |
| Birth type | | | | | $\chi^2(6) = 25.73$, ***<br>V=.06 |
| Spontaneous | 1124 (66.7%)<sup>a</sup> | 1089 (64.4%)<sup>a,b</sup> | 1392 (61.5%)<sup>b,c</sup> | 106 (54.6%)<sup>c</sup> |  |
| Cesarean | 413 (24.5%)<sup>a</sup> | 465 (27.5%)<sup>a,b</sup> | 673 (29.7%)<sup>b</sup> | 76 (39.2%)<sup>c</sup> |  |
| Instrumental | 129 (7.7%)<sup>a</sup> | 120 (7.1%)<sup>a</sup> | 175 (7.7%)<sup>a</sup> | 11 (5.7%)<sup>a</sup> |  |
| Missing | 20 (1.2%) | 17 (1.0%) | 25 (1.1%) | 1 (0.005%) |  |
| Multiple birth | | | | | $\chi^2(3) = 9.41$, ***<br>V=.04 |
| Single | 1602 (95.0%)<sup>a</sup> | 1590 (94.0%)<sup>a,b</sup> | 2131 (94.1%)<sup>a,b</sup> | 174 (89.7%)<sup>b</sup> |  |
| Twin or more | 84 (5.0%)<sup>a</sup> | 101 (6.0%)<sup>a,b</sup> | 134 (5.9%)<sup>a,b</sup> | 20 (10.3%)<sup>b</sup> |  |
| Mother's age, mean<br>(SD, Min-Max) | 31.48 (5.83, 16-54)<sup>a</sup> | 32.2 (5.96, 17-51)<sup>b</sup> | 31.54 (6.03, 18-53)<sup>a</sup> | 31.55 (5.69, 20-47)<sup>a,b</sup> | F (3,5812)<br>=5.41***, $\eta^2$<br>=.003 |
| Missing | 10 (0.005%) | 10 (0.005%) | 4 (0.001%) | 0 |  |
| Child nationality | | | | | $\chi^2(3) = 106.88$,<br>*** V=.14 |
| Jewish | 1209 (71.7%)<sup>a</sup> | 1172 (69.3%)<sup>b</sup> | 1343 (59.3%)<sup>c</sup> | 136 (70.1%)<sup>a,b</sup> |  |
| Other | 265 (15.7%)<sup>a</sup> | 337 (19.9%)<sup>b</sup> | 651 (28.7%)<sup>c</sup> | 40 (20.6%)<sup>a,b</sup> |  |
| Missing | 212 (12.6%) | 182 (10.8%) | 271 (12.0%) | 18 (9.3%) |  |
| Mother birth country | | | | | $\chi^2(3) = 82.8$, ***<br>V=.12 |
| Israel | 1289 (76.5%)<sup>a</sup> | 1203 (71.1%)<sup>b</sup> | 1455 (64.2%)<sup>c</sup> | 157 (80.9%)<sup>a</sup> |  |
| Other | 291 (17.3%)<sup>a</sup> | 355 (21.0%)<sup>b</sup> | 628 (27.7%)<sup>c</sup> | 24 (12.4%)<sup>a</sup> |  |
| Missing | 106 (6.3%) | 133 (7.9%) | 182 (8.0%) | 13 (6.7%) |  |

| SES status | | | | | $\chi^2(6) = 141.65$,<br>*** $V = .12$ |
| --- | --- | --- | --- | --- | --- |
| Low | 185 (11%)<sup>a</sup> | 135 (8%)<sup>a</sup> | 203 (9%)<sup>a</sup> | 21 (10.8%)<sup>a</sup> |  |
| Low- medium | 367 (21.8%)<sup>a</sup> | 446 (26.4%)<sup>b</sup> | 692(30.6%)<sup>c</sup> | 63(32.5%)<sup>b,c</sup> |  |
| Medium- high | 448 (26.6%)<sup>a</sup> | 457 (27%)<sup>a</sup> | 570(25.2%)<sup>a</sup> | 46(23.7%)<sup>a</sup> |  |
| High | 340 (20.2%)<sup>a</sup> | 225 (13.3%)<sup>b</sup> | 174 (7.7%)<sup>c</sup> | 7 (3.6%)<sup>c</sup> |  |
| Missing | 346 (20.5%) | 428 (25.3%) | 626 (27.6%) | 57 (29.4%) |  |

*Note.* Missing values were not included in the above analyses. Due to multiple comparisons, Bonferroni correction was applied, yielding a p-value threshold of $p < .001$ . For nominal variables, the letters represent significant cluster differences based on adjusted pairwise z-tests. For continuous variables, the letters represent significant cluster differences based on Bonferroni post hoc tests. \*\*\* $p < .001$

Children in the severe cluster had significantly lower birth weights and higher rates of preterm, multiple births, and cesarean births relative to the other EDM clusters, which did not differ (Table 2).

**Table 2.** Rates of Parent and Provider Concerns by EDM Cluster.

| Characteristic | Typical (n=1,686) | Mild (n=1,691) | Moderate (n=2,265) | Severe (n=194) | Statistics |
| --- | --- | --- | --- | --- | --- |
| Parent concern for development | 486 (28.8%)<sup>a</sup> | 811 (48.0%)<sup>b</sup> | 1465 (64.7%)<sup>c</sup> | 162 (83.5%)<sup>d</sup> | $\chi^2(3) = 587.52$,<br>*** $V = .31$ |
| Parent hearing suspicion | 146 (8.7%)<sup>a</sup> | 247 (14.6%)<sup>b</sup> | 539 (23.8%)<sup>c</sup> | 77 (39.7%)<sup>d</sup> | $\chi^2(3) = 231.37$,<br>*** $V = .19$ |
| Developmental referral | 64 (3.8%)<sup>a</sup> | 136 (8%)<sup>b</sup> | 247 (10.9%)<sup>c</sup> | 24 (12.4%)<sup>b,c</sup> | $\chi^2(3) = 70.7$,<br>*** $V = .11$ |
| Missing | 10 (0.6%) | 6 (0.4%) | 11 (0.5%) | 0 |  |
| Number of visits, mean (SD, Min-Max) | 13.52 (3.05, 7-22) | 13.56 (2.87, 8-22) | 13.4 (2.83, 7-22) | 13.35 (3.12, 8-22) | F (3,5684) = 1.29, $\eta^2 = .001$ |
| Missing | 50 (0.02%) | 38 (0.02%) | 50 (0.02%) | 14 (0.07%) |  |

*Note.* Missing values were not included in the above analyses. Due to multiple comparisons Bonferroni correction was applied, yielding a p-value threshold of $p < .001$ . Different letters represent significant cluster differences based on adjusted pairwise z-tests.
\*\*\* $p < .001$

### Descriptive

For the severe cluster, milestone passing rates in the motor domains were lower and declined over time. Rates dropped below 50% from the 9–12 month visit. The moderate cluster showed lower rates of passing for later fine motor milestones such as 70.02% 18–24 months ‘building tower with cubes’ and 46.13% 24–36 months ‘imitating lines’. Note that the mild cluster had around 85% passing rate for later motor milestones, particularly fine motor (See Figure 1).

In the language and social domains, the severe cluster had lower passing rates during the 9–12 month visit with 71.64% “understand simple instructions” and 71.65% “says one word or pronounces meaningful sounds” to almost no child passing beyond the 24 month point. The moderate cluster was next in passing rates. They decreased from 91.39% of children passing the 12-18 months milestone “express vocally or with gestures” to 40% passing the 18-24 months milestone of “has a vocabulary of over ten words” to not passing at all by 36 months. Next was the mild cluster which had 70–80% passing rates at 18–24 months “composes two word sentences” to under 10% by 36 months (See Figure 1; See supplemental Table 1 for a summary of EDM cluster differences).

Figure 2 presents the mean percentage of failed domain milestones by EDM clusters. Language differentiated all four EDM clusters. The severe cluster was distinct in its higher percentage of failed milestones relative to the other clusters across domains but was particularly high in the gross motor and language domains. The 3 children who failed all milestones in the gross motor and language domains were from the severe cluster.

**Figure 2.**
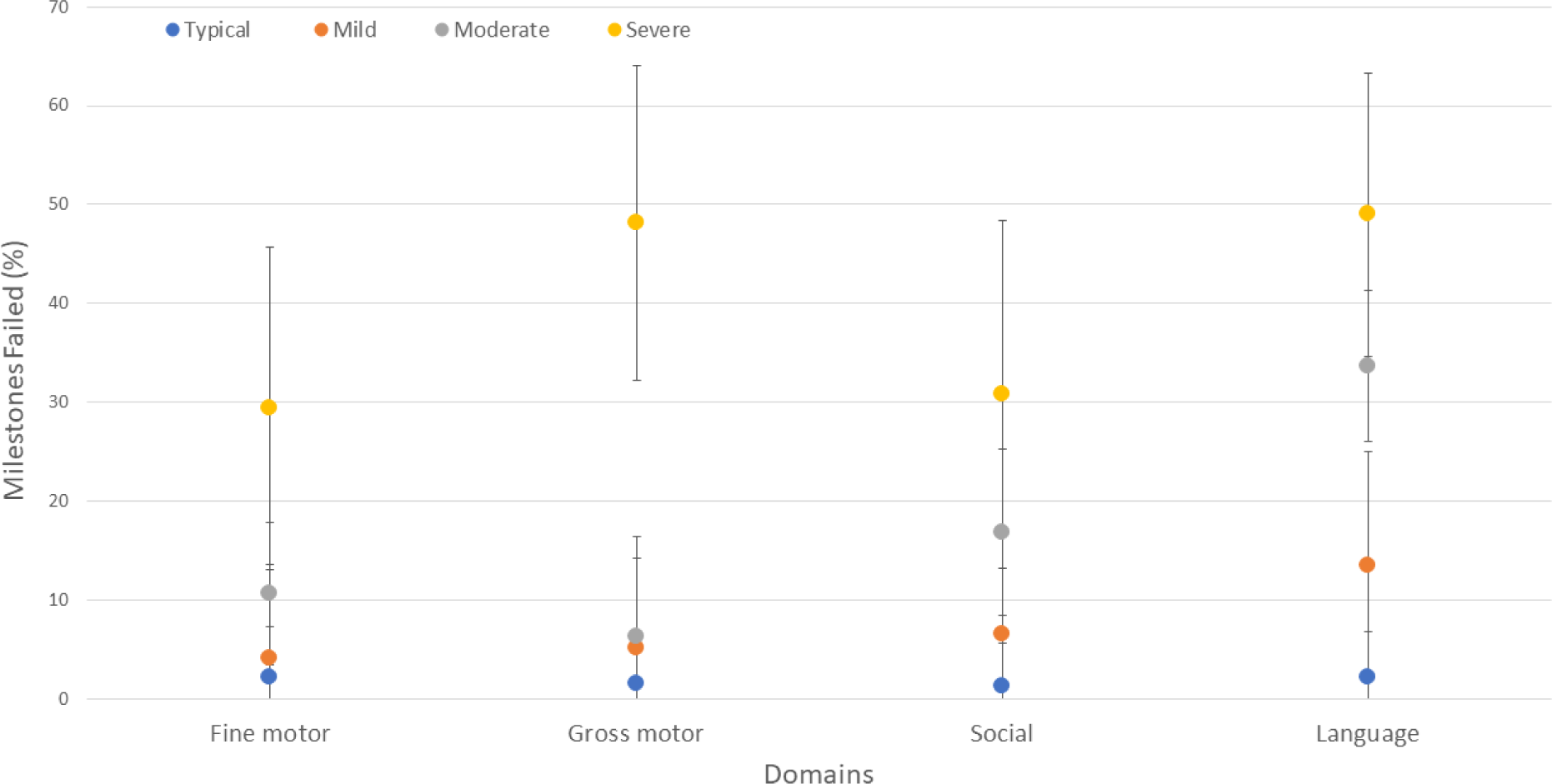
Mean (SE) number of failed milestones within each domain by EDM cluster. *Note.* There are 15 language, 7 social, 10 fine motor and 11 gross motor milestones.

Figure 3 displays mean domain percentiles age at first performance over time. Mean percentiles of typical, mild, and moderate EDM clusters in the motor domains ranged from 0.47–0.55, and were relatively stable. These percentiles did not overlap with the range of percentiles in the severe cluster (0.29–0.40). The severe cluster showed a dip 12-24 months in gross and fine motor percentiles and improved by 24-36 months. In the social domain, the severe cluster started lower but reached close to the other clusters by 36 months. Typical, mild and moderate clusters’ social percentiles ranged from 0.46–0.54 across time. In the language percentile, the severe cluster decreased by about 12 points over three years, while the other clusters decreased by about 5 percentiles.

**Figure 3.**
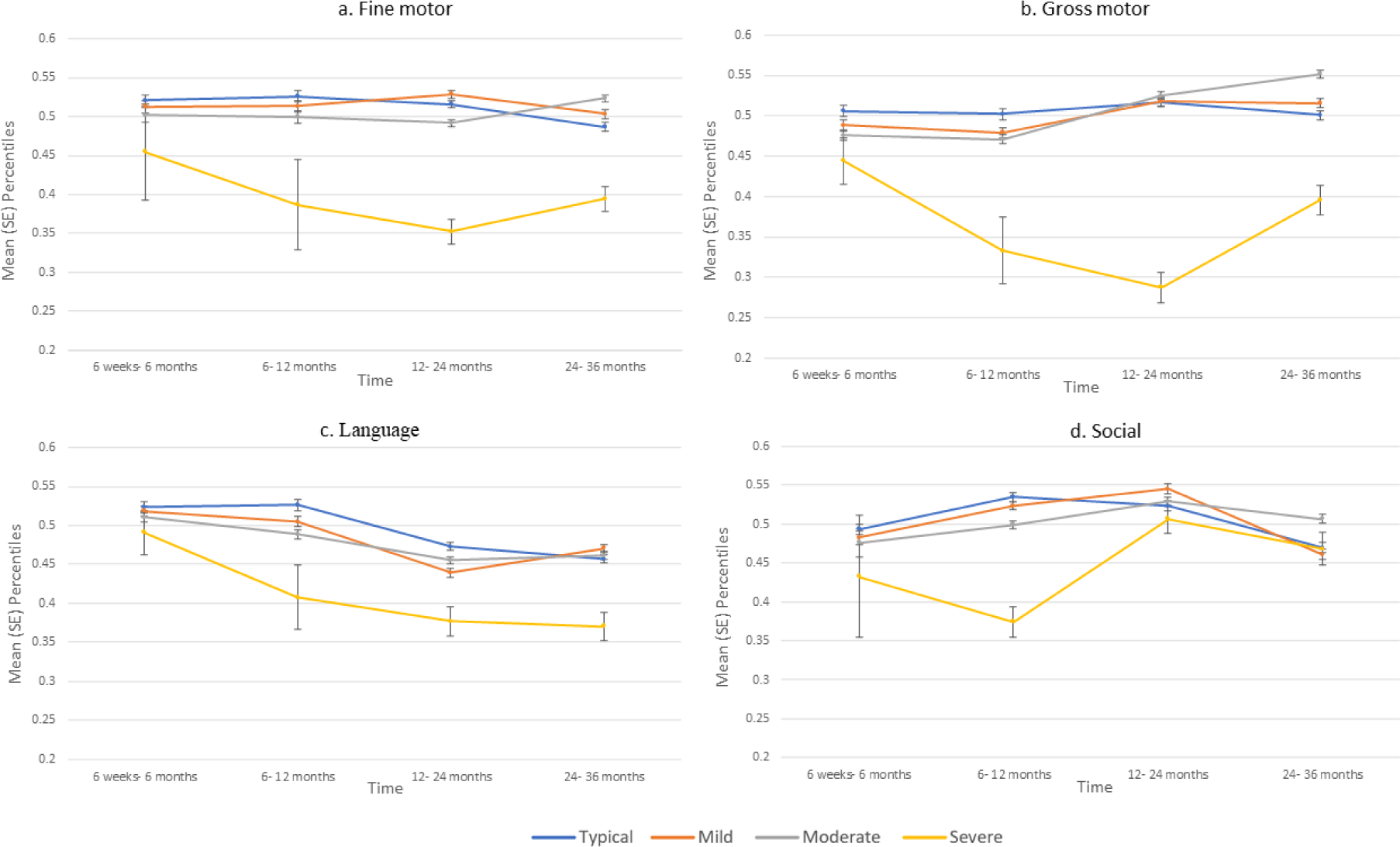
a-d. Mean (SE) domain percentiles age at first performance across times by EDM cluster *Note.* Each of the first three age ranges presented group data from two visits so that 6 weeks to 6 months.

### Developmental validation of EDM clusters

Chi-square tests indicated that all four clusters significantly differed in the rates of parental concerns for development, parental suspicion of hearing impairment, and provider developmental referral (Table 2). Cramer’s V showed medium to large effect sizes for these variables. Results for all three variables validate the ranking of clusters from the most concerns and referrals to the least as follows: severe, moderate, mild and typical. The number of baby wellness visits was not significantly different between clusters.

## Discussion

ASD Children often exhibit developmental delays during the first two years of life [22,24]. However, it is essential to recognize that there is considerable variability in the developmental paths of ASD children, challenging early identification and timely intervention [5,13]. This birth cohort study is unique in its approach to clustering ASD children based on a comprehensive set of developmental milestones routinely recorded prospectively and sequentially from 0–36-months, by healthcare providers. These EDM clusters differed in birth parameters, demographic features and rates of parental concerns and provider referral. The largest cluster, comprising 38.81% of the cohort, exhibited moderate language and social delays prominent in the second and third years-of-age with later fine motor delays. This cluster had the highest rate of immigrant and non-Jewish mothers. Next was a cluster with mild language delays (28.98%) prominent in the third year of life and more immigrant mothers, and a cluster generally intact developmentally and higher socioeconomically (28.89%). The smallest cluster (3.3%) presented greater severity overall and early gross motor delays, higher rate of preterm and cesarean birth and lower socioeconomically. These diverse developmental profiles highlight variations in the severity, onset, and domains of delays among ASD children, defining four distinct early endophenotypes. Understanding these differences is crucial for correlating phenotype to genotype or other etiologies, and tailoring intervention strategies to address the unique needs of each subgroup.

### Specific trajectories identified through each EDM cluster

The severe EDM cluster displayed the most pronounced delays across developmental domains, falling behind by an average of 30%–50% of domain milestones. Notably, this cluster exhibited the lowest percentiles for age at first performed gross motor, fine motor, and language milestones, with no overlap with the other clusters. The considerable variance in developmental scores within the severe cluster can be attributed, in part, to its smaller size. At the individual milestone level, signs of deviation within the severe cluster began to manifest during the 6–9-month visit, with failure rates exceeding 10% for motor milestones such as “transferring an object between hands,” and language-social milestones like “saying repetitive syllables.” By the 24–36-month assessment, this cluster exhibited failure rates of 90%-100% across all milestones. Additionally, this cluster had significantly higher rates of preterm and cesarean births and lower birth weights, indicating a more complex medical profile. This link between motor delays and prematurity suggests that this developmental trajectory may provide information on additional etiologies linked to ASD, such as Fragile X syndrome [13]. The small size of the severe cluster may reflect an inherent challenge in early ASD diagnosis of infants with intricate medical and developmental profiles. In the presence of early motor delays, it is imperative to closely monitor communication development.

The age at which passing rates declined differed between EDM clusters and developmental domains. For language milestones, the severe cluster exhibited a gradual drop in passing rates starting at 9 months and falling below 50% by the 12–18-month visit. In contrast, the moderate cluster experienced a large drop in passing rates at the 18–24-month visit, while the mild cluster displayed a decline at the 24–36-month visit. Concerning motor milestones, the severe cluster began to show declining passing rates as early as the 6–9-month visit, falling below 50% by the 12–18-month visit. The passing rates of the moderate cluster declined below 50% only at the 24–36-month visit, while the other clusters maintained passing rates above 80% at all visits. These findings align with prior research, of a severe ASD cluster displaying early motor delays at 6 months and the typical cluster primarily manifesting social-communication delays in the second year of life [30]. The moderate cluster primarily experienced later fine motor delays compared to the early gross motor delays of the severe cluster.

Studies following the same protocol used either percent of pass/fail within an age range in the general population [28] or pass/fail in ASD [22,24]. Using pass/fail captures the progress of all children without indicating degree of delay, while percentiles based on age at passing showed the magnitude of delay but only captures children who pass. We used three methods to extract meaning from this clinical protocol: 1. Pass/fail (figure 1); 2. Percent of failed milestones within domain (figure 2); and 3. Mean domain percentiles age at passing at 4 time points (figure 3). The findings highlight how these approaches complement each other. Typical, moderate, severe clusters presented language delays during second and third years of life, as presented by mean percentiles below 50. The typical cluster showed a mild language delay in the third year. Those who pass do not necessarily pass on time and at the first testing attempt. We see greater variability between and within EDM clusters in the percent who pass a milestone. Looking at mean language percentiles, the mild, moderate and typical clusters were relatively similar; whereas the percentage of failed language milestones differentiated all four EDM clusters. It appears that those in the moderate cluster who pass are mildly delayed; however, many fail and thus are not captured in the percentile findings.

### EDM Cluster validation

The EDM clusters’ differences were validated by differences in the percentage of parental concerns and provider referrals among them. Notably, the severe cluster exhibited the highest frequency of both developmental concerns and referrals, followed by the moderate cluster. In contrast, the typical and the mild clusters displayed lower rates of developmental concerns and referrals. This order corresponds to the magnitude as well as the onset of their delay. Although not a core ASD symptom, language delays are among the most common first concerns of parents of ASD children [31,32]. The present study adds valuable insights into the connection between parental concerns and phenotypic variations because parental concern was part of a prospective developmental screening. Future research endeavors aimed at exploring the connection between cluster dependency and the age of diagnosis may untangle the influence of a particular developmental profile on early detection and subsequent trajectory.

In the severe cluster, the heightened level of concern regarding development can be attributed to the increased awareness among parents and professionals regarding motor milestones during infancy, as these are typically easier to recognize. Conversely, language development, which tends to raise concerns more prominently after the age of two, is often less conspicuous early on. This supports prior studies suggesting a link between parental concern, motor delays and age of autism detection [31]. For instance, research shows that parental concerns regarding motor development predict ASD as early as 6 months, while parental concerns regarding communication predict ASD only after 12 months [32]. While evidence shows that only 12.5% of parent’s first concerns for ASD children relate to motor delays, these concerns appear at an earlier age compared to concerns related to language delay [31]. Furthermore, ASD children are referred earlier for motor delays versus language and communication delays [21]. The later fine motor delays of the moderate cluster may have also contributed to their higher concerns and referrals.

### Socioeconomics and ASD development

Variability in early development is closely linked to socioeconomic factors. The moderate and mild EDM clusters, which have greater language compared to motor impairments, included more non-Jewish and immigrant mothers. In contrast, the typical cluster had the lowest rates of non-Jewish, immigrant mothers and the highest percentage of high socioeconomic status; revealing how developmental trajectories are related to family and community demographics. While prior Israeli ASD research did not find an association between ethnicity and milestone failure [22], our findings highlight the role of ethnicity and immigration status in explaining within-group variability. In the USA, representative studies show lower ASD prevalence and delayed diagnosis in minority groups, but once diagnosed lower IQ levels [33]. These minority groups also display a non-classic clinical phenotype characterized by heightened emotional dysregulation [34]. One hypothesis suggests that minority children may need to surpass a higher threshold of difficulties to obtain an ASD diagnosis, possibly due to provider bias in recognizing symptoms in non-native speaking families and a greater threshold for parents seeking help [35]. However, family minority status seemed less relevant for the severe group, suggesting a more complex neurogenetic etiology that minimizes the impact of environmental factors on diagnostic thresholds.

## Limitations

Classification included only ASD children with complete milestone data (36.58%) to enable meaningful interpretation of common profiles. Encouraging were the similar rates of complete data in typically developing children. However, this may have created bias by excluding children who are more strongly affected, as they are less likely to show up for later wellness visits due to receiving developmental therapy elsewhere. Future research should focus on understanding which milestones are critical for differentiating clusters. Clinically, adding techniques to increase milestone data collection from all children is warranted.

The developmental protocol analyzed has several limitations to consider in interpreting the results. First, the milestones are not distributed equally across domains and visits. The observation that up to 6 months most ASD children in this cohort passed milestones may reflect the nature of the screening protocol, as the first 6 months contains fewer social and language items than in later visits. Social milestones didn’t distinguish the four clusters as well as the language ones, suggesting the need for refining this domain specifically for ASD detection. In addition, ASD children, and the more motorically-affected clusters in particular, had more difficulty passing later fine motor milestones such as ‘building a tower’, and ‘imitating lines’. Typically developing children attained the fine motor milestones in this protocol earlier than the protocol’s expected age, with little variance [28]. These milestones also require language and social cognition; thus, may reflect difficulties beyond fine motor skills. Motor evaluation that does not depend on social-communication abilities is needed [36].

Further research is needed to reveal the relation between distinct early developmental profiles with age of diagnosis, medical comorbidities, genetic etiology and intervention outcomes. Age of ASD diagnosis and severity were missing in the database. We hypothesize that the severe cluster was diagnosed earlier and has greater ASD severity and that the typical cluster is linked with later diagnosis and less support. The differences in age and type of delay may correspond with medical comorbidities, as reported previously [10]. In addition, patterns of improvement in percentiles between the second and third years of life can reflect intervention outcomes following diagnosis but may also reflect the complex nature of the later milestones. Previous research showed that most ASD children achieved significant gains in language and communication skills, with the most severe cluster showing the least improvement [25]. The nature and course of delays in each EDM cluster calls for a personalized intervention approach at the prodromal phase considering both birth parameters and sociodemographic moderators.

## Conclusions

Across four identified EDM clusters, disparities in the extent of delays in each domain and their evolution over time were observed, separating into four distinct trajectories. While all EDM clusters showed some degree of delay, they differed in magnitude and onset. Children in the most severe cluster showed signs in the first year of life, the moderate in the second year, the language in the third year, whereas the typical cluster passed most milestones. Motor delays were associated with earlier concerns and referrals. In addition, the findings underscore the interplay of sociodemographic and birth-related factors with clusters’ developmental trajectories, highlighting that early detection and progress are shaped by a complex interplay of neuro-genetic and environmental variables. Recognizing early developmental patterns across domains and over time can inform further investigations into distinct subgroups of ASD children with specific developmental requirements, potentially aligning with intervention needs and etiology (e.g., prematurity and genetic syndromes). These unique early developmental profiles offer valuable insights for early screening efforts, enhancing provider awareness of the early developmental variability, as evident in the analysis of developmental surveillance data routinely collected by healthcare providers.

## Data Availability

All data studied is confidential and belongs to the Israeli Ministry of Health.

## References

1. American Psychiatric Association. Diagnostic and statistical manual of mental disorders. 5th ed. Washington D.C. 2013

2. Jacob S, Wolff JJ, Steinbach MS, Doyle CB, Kumar V, Elison JT. Neurodevelopmental heterogeneity and computational approaches for understanding autism. Transl psychiatry. 2019; 9(1):63.

3. Stevens E, Dixon DR, Novack MN, et al. Identification and analysis of behavioral phenotypes in autism spectrum disorder via unsupervised machine learning. Int J Med Inform. 2019;129:29–36. DOI: 10.1016/j.ijmedinf.2019.05.006.

4. Syriopoulou-Delli CK, Papaefstathiou E. Review of cluster analysis of phenotypic data in Autism Spectrum Disorders: distinct subtypes or a severity gradient model?. Int J of Dev Disabil. 2020;66(1):13–21. DOI: 10.1080/20473869.2018.1542561.

5. Wiggins LD, Tian LH, Lev SE et al. Homogeneous subgroups of young children with autism improve phenotypic characterization in the study to explore early development. J Autism Dev Disord. 2017;47:3634–3645. DOI: 10.1007/s10803-017-3280-4.

6. Beversdorf DQ. Phenotyping, etiological factors, and biomarkers: toward precision medicine in autism spectrum disorders. J Dev Behav Pediatr. 2016;37(8):659–673. DOI: 10.1097/DBP.0000000000000351.

7. Arnett AB, Beighley JS, Kurtz-Nelson EC, et al. Developmental predictors of cognitive and adaptive outcomes in genetic subtypes of autism spectrum disorder. Autism Res. 2020;13(10):1659–1669. DOI: 10.1002/aur.2385.

8. Lord C, Bishop S, Anderson D. Developmental trajectories as autism phenotypes. Am J Med Genet C Semin Med Genet. 2015;169(2):198–208. DOI: 10.1002/ajmg.c.31440.

9. Zheng S, Hume KA, Able H, Bishop SL, Boyd BA. Exploring developmental and behavioral heterogeneity among preschoolers with ASD: A cluster analysis on principal components. Autism Res. 2020;13(5), 796–809.

10. Doshi-Velez F, Ge Y, Kohane I. Comorbidity clusters in autism spectrum disorders: an electronic health record time-series analysis. Pediatrics. 2014;133(1):e54–e63. doi:10.1542/peds.2013-0819

11. Montgomery A, Masi A, Whitehouse A, et al. Identification of subgroups of children in the Australian Autism Biobank using latent class analysis. Child Adolesc Psychiatry Ment Health. 2023;17(1):1–12. DOI: 10.1186/s13034-023-00565-3.

12. Vargason T, Frye RE, McGuinness DL, Hahn J. Clustering of co-occurring conditions in autism spectrum disorder during early childhood: A retrospective analysis of medical claims data. Autism Res. 2019;12(8):1272–1285. DOI: 10.1002/aur.2128.

13. Kuo SS, van der Merwe C, Fu JM, et al. Developmental variability in autism across 17 000 autistic individuals and 4000 siblings without an autism diagnosis: comparisons by cohort, intellectual disability, genetic etiology, and age at diagnosis. JAMA pediatr. 2022;176(9):915–923. DOI: 10.1001/jamapediatrics.2022.2423.

14. Corbett BA, Schwartzman JM, Libsack EJ, et al. Camouflaging in Autism: Examining Sex-Based and Compensatory Models in Social Cognition and Communication. Autism Res. 2021;14(1):127–142. DOI: 10.1002/aur.2440. Epub 2020 Nov 21.

15. Kentrou V, de Veld DM, Mataw KJ, Begeer S. Delayed autism spectrum disorder recognition in children and adolescents previously diagnosed with attention-deficit/hyperactivity disorder. Autism. 2019;23(4):1065–1072. doi: 10.1177/1362361318785171.

16. Lombardo MV, Lai MC, Baron-Cohen S. Big data approaches to decomposing heterogeneity across the autism spectrum. Mol psychiatry. 2019;24(10):1435–1450.

17. Ozonoff S, Heung K, Byrd R, Hansen R, Hertz-Picciotto I. The onset of autism: patterns of symptom emergence in the first years of life. Autism Res. 2008;1(6):320–328. DOI: 10.1002/aur.53.

18. Wozniak RH, Leezenbaum NB, Northrup JB, West KL, Iverson JM. The development of autism spectrum disorders: variability and causal complexity. Wiley Interdiscip Rev Cogn Sci, 2017;8(1-2), e1426. DOI: 10.1002/wcs.1426.

19. Yates K, Le Couteur A. Diagnosing autism/autism spectrum disorders. Paediatr Child Health. 2016;26(12):513–518. DOI: 10.1016/j.paed.2016.08.004.

20. Fountain C, Winter AS, Bearman PS. Six developmental trajectories characterize children with autism. Pediatrics. 2012;129(5):e1112–e1120. DOI: 10.1542/peds.2011-1601.

21. Gurevitz M, Leisman G. Factors in Infancy That May Predict Autism Spectrum Disorder. Brain Sci. 2023;13(10):1374. DOI: 10.3390/brainsci13101374.

22. Alhozyel E, Elbedour L, Balaum R, et al. Association between early developmental milestones and autism spectrum disorder. Res Child Adolesc Psychopathol. 2023;1–10. DOI: 10.1007/s10802-023-01085-6.

23. Shan L, Feng JY, Wang TT, Xu ZD, Jia FY. Prevalence and Developmental Profiles of Autism Spectrum Disorders in Children With Global Developmental Delay. Front Psychiatry. 2022;12:794238. DOI: 10.3389/fpsyt.2021.794238.

24. Davidovitch M, Stein N, Koren G, Friedman BC. Deviations from typical developmental trajectories detectable at 9 months of age in low risk children later diagnosed with autism spectrum disorder. J Autism Dev Disord. 2018;48(8):2854–2869. DOI: 10.1007/s10803-018-3549-2.

25. Kim SH, Macari S, Koller J, Chawarska K. Examining the phenotypic heterogeneity of early autism spectrum disorder: subtypes and short-term outcomes. J Child Psychol Psychiatry. 2016;57(1):93–102. DOI: 10.1111/jcpp.12448.

26. Rosello R, Berenguer C, Martinez-Raga J, Miranda A, Cortese S. Subgroups of Children with Autism Spectrum Disorder without Intellectual Disability: A Longitudinal Examination of Executive and Socio-Adaptive Behaviors in Adolescence. J Clin Med. 2021; 10(10):2220. DOI: 10.3390/jcm10102220

27. Mosconi MW, Stevens CJ, Unruh KE, Shafer R, Elison JT. Endophenotype trait domains for advancing gene discovery in autism spectrum disorder. J Neurodevelop Dis. 2023;15(1):1–27. DOI: 10.1186/s11689-023-09511-y.

28. Sudry T, Zimmerman DR, Yardeni H, et al. Standardization of a developmental milestone scale using data from children in Israel. JAMA netw open. 2022;5(3):e222184–e222184. DOI: 10.1001/jamanetworkopen.2022.2184.

29. Frankenburg WK, Dodds JB. The Denver developmental screening test. J Pediatr. 1967.

30. Estes A, Zwaigenbaum L, Gu H, et al. Behavioral, cognitive, and adaptive development in infants with autism spectrum disorder in the first 2 years of life. J Neurode. Disord. 2015;7:1–10.

31. Guinchat V, Chamak B, Bonniau B, et al. Very early signs of autism reported by parents include many concerns not specific to autism criteria. Res Autism Spectr Disord. 2012;6(2):589–601.

32. Sacrey LAR, Zwaigenbaum L, Bryson S, et al. Can parents’ concerns predict autism spectrum disorder? A prospective study of high-risk siblings from 6 to 36 months of age. J Am Acad Child Adolesc Psychiatry. 2015;54(6):470–478. DOI: 10.1016/j.jaac.2015.03.014.

33. Baio J, Wiggins L, Christensen DL, et al. Prevalence of autism spectrum disorder among children aged 8 years—autism and developmental disabilities monitoring network, 11 sites, United States, 2014. MMWR Surveill Summ. 2018;67(6):1. DOI: 10.15585/mmwr.ss6706a1.

34. Becerra TA, von Ehrenstein OS, Heck JE, et al. Autism spectrum disorders and race, ethnicity, and nativity: a population-based study. Pediatrics. 2014;134(1):e63–e71. DOI: 10.1542/peds.2013-3928.

35. Angell AM, Empey A, Zuckerman KE. A review of diagnosis and service disparities among children with autism from racial and ethnic minority groups in the United States. INT REV RES DEV DISA. 2018;55:145–180.

36. Wilson RB, McCracken JT, Rinehart NJ, Jeste SS. What’s missing in autism spectrum disorder motor assessments?. J. Neurodev. Disord. 2018;10(1):1–13.

