## Supplemental Materials for "Early developmental milestone clusters of autistic children based on electronic health records"

Figure S1. Distribution of the number of missing milestones among ASD children

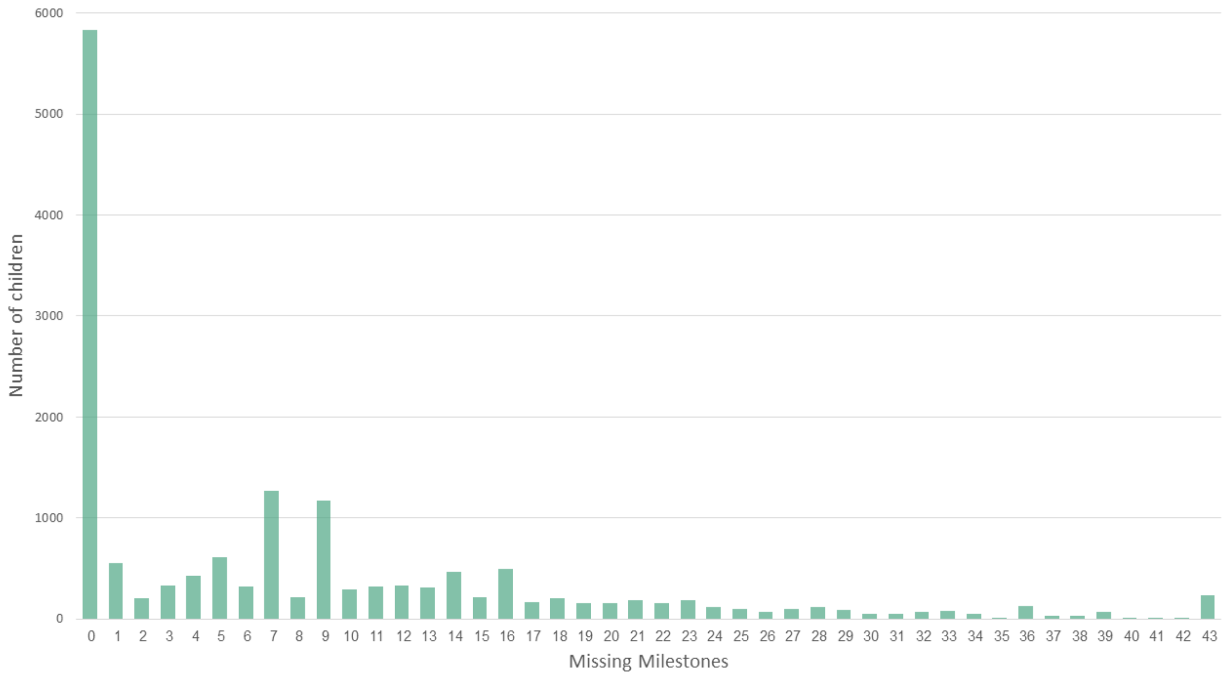


*Note.* 5836 ASD children had 0 missing milestones.

Figure S2. The number of participants in each of 9 clusters.

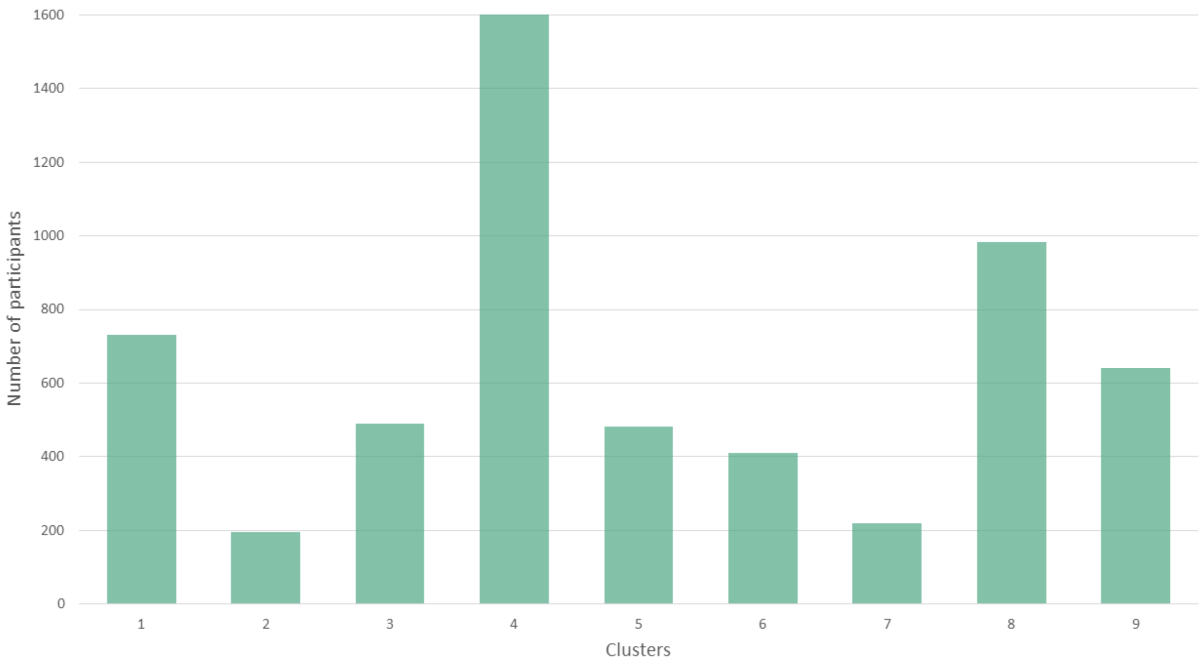

Figure S3 Development of clusters across time in language-social relative to motor domains.

3a. Development of initial 9 clusters across time.

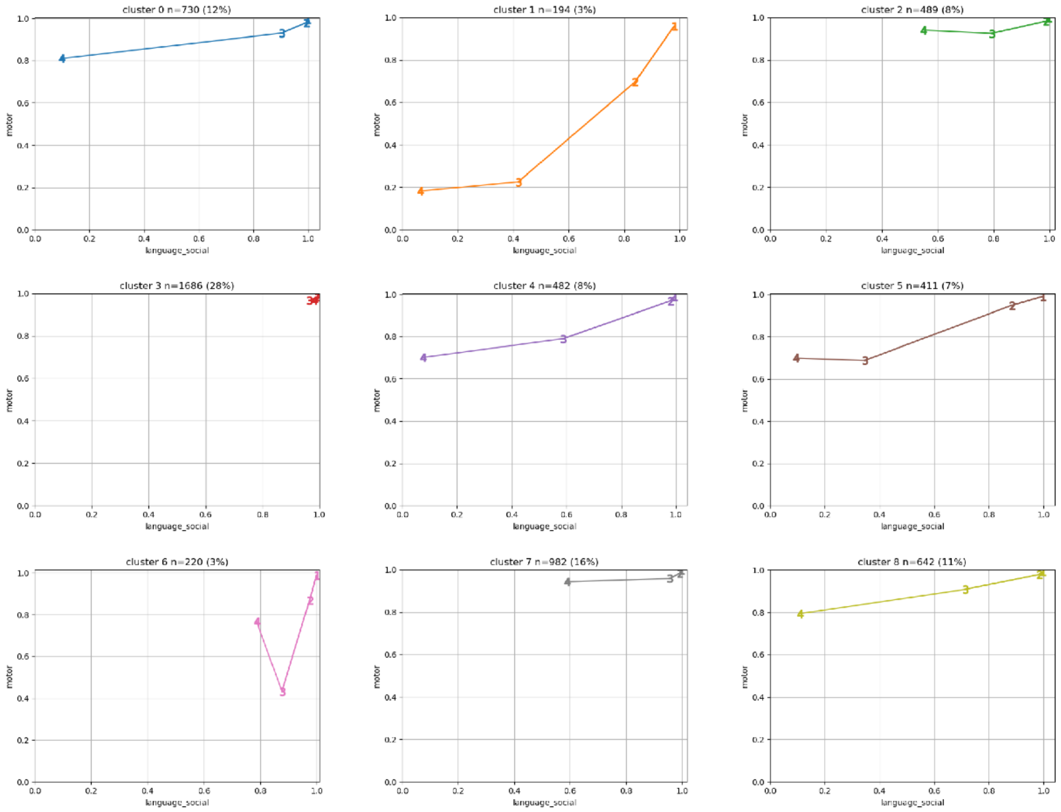


*Note.* For descriptive purposes, mean passing rates in each domain were computed for 4 time points: time 1= up to 6 months, time 2= 6–12 months, time 3=12–24 months, time 4=24–36 months. For visualization we grouped fine and gross motor into the motor axis, and language and social milestones into a language-social axis. These nine clusters were grouped into four clusters based on their similarities as follows: Typical cluster= cluster 3 high across all points. Moderate cluster= clusters 0,4,5,8 with a mixed language-social and mild motor involvement. Mild cluster= clusters 2,6,7 language-social delay. Severe cluster=cluster 1 with low language-social scores as motor delay.

3b. Development of 4 clusters across time.

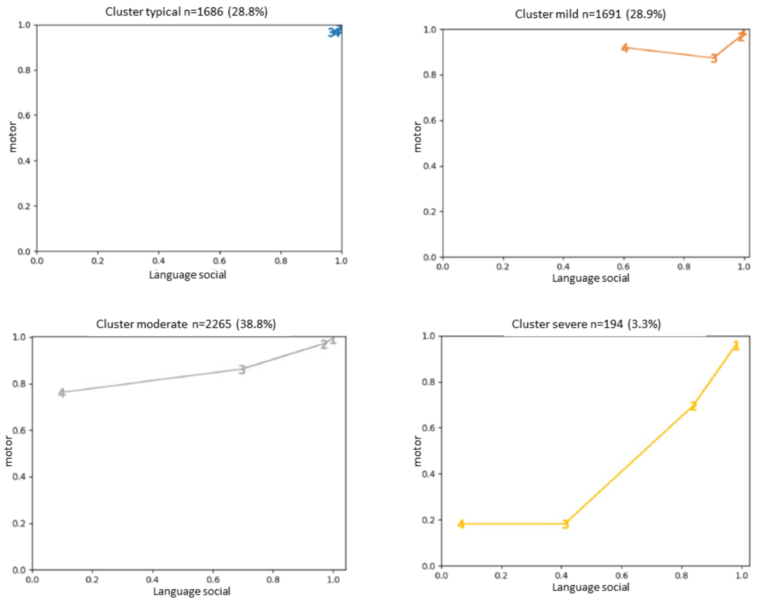


*Note.* Numbers on graph points represent times. Motor refers to gross and fine motor and language_social refers to language and social domains. Time 1= up to 6 months, time 2= 6-12 months, time 3=12-24 months, time 4=24-36 months.

Table S1. Summary of Common Features for EDM Clusters

| | *Milestone Delay (months)* | | | | | |
|---|---|---|---|---|---|---|
| | ***Language / Social*** | ***Fine Motor*** | ***Gross Motor*** | ***Birth Parameters*** | ***Demo-graphics*** | ***Parent concerns*** |
| EDM Typical | No delay | No delay | No delay | | High SES | Low |
| EDM Mild | **24-36** | No delay | No delay | | Medium SES | Medium |
| EDM Moderate | **12-18** greater magnitude from **18** months | **18-36** | No delay | | Low SES | High |
| EDM Severe | **9-12** greater magnitude from **12** months. | **6-9** | **6-9** | Low birth weight. Preterm Cesarean birth | Low SES | Highest |
